# Edge-Based ADL Recognition Using Room-Specialized Mixture-of-Experts

**DOI:** 10.64898/2026.07.24.26357042

**Authors:** Venkatanand ram Addepalli, Praveen Rao, Knoo Lee

## Abstract

Activities of daily living (ADLs) provide important indicators of functional decline in people living with dementia, motivating the need for continuous in-home monitoring. However, deploying transformer-based activity recognition models on resource-constrained edge devices remains challenging because of limited computational resources and the need to preserve participant privacy by avoiding cloud-based processing. In this work, we propose a room-specialized Mixture-of-Experts (MoE) architecture for edge-based ADL recognition using ambient smart home sensors. Household activities are decomposed into room-specific transformer experts through deterministic routing, while temporal subsampling bounds the computational cost of each activity segment, enabling efficient on-device learning and inference. We evaluated the proposed framework using data collected from five dementia households, achieving Macro-F1 scores ranging from 0.437 to 0.911 despite substantial differences in activity distributions across homes. End-to-end training was successfully performed on a Raspberry Pi, demonstrating the feasibility of transformer-based ADL recognition on low-cost edge hardware. These findings suggest that room-specialized MoE provides a practical, privacy-preserving framework for continuous smart home monitoring and establishes a foundation for future edge-native healthcare applications, including continual and federated learning.

## I. Introduction

Alzheimer’s disease is the most common cause of dementia, yet timely assessment remains challenging because of workforce shortages and prolonged wait times for clinical evaluation [1, 2]. Functional changes in activities of daily living (ADLs) often emerge early in disease progression, making continuous, in-home monitoring ideal complement to episodic clinical assessments.

In the United States, an estimated 7.4 million Americans aged 65 and older were living with Alzheimer’s dementia in 2026[1]. As dementia progresses, functional impairments extend from activities of daily living (ADLs) to instrumental activities of daily living (IADLs), increasing dependence on caregivers and reducing independent living.. Nearly 13 million unpaid family caregivers provide most of this support [1]. These progressive functional changes highlight the need for objective, continuous assessment of everyday functioning outside traditional clinic visits, where subtle changes in daily behavior may otherwise go undetected.

Deploying continuous activity recognition in real homes requires artificial intelligence models that can operate directly on resource-constrained edge devices, avoiding reliance on cloud infrastructure while preserving participant privacy. However, deploying transformer-based activity recognition models on edge hardware remains challenging because of limited computational resources and memory available on devices such as the Raspberry Pi[3]. Rather than asking whether edge devices can match workstation-level performance, we ask whether transformer-based ADL recognition can be redesigned to operate efficiently within these resource constraints. Household activities naturally occur within distinct rooms, resulting in localized sensor patterns that can be modeled independently rather than by a single global model [4]. Motivated by this observation, we propose a room-specialized Mixture-of-Experts architecture in which lightweight transformer experts are assigned to individual rooms through deterministic routing. Combined with temporal subsampling of activity segments, the proposed framework bounds computational cost while enabling end-to-end training and inference on Raspberry Pi devices [5].

The main contribution of this work are as follows: A room-specialized Mixture-of-Experts architecture for edge-based ADL recognition; deterministic room-based routing with temporal subsampling for efficient on-device transformer deployment; and end-to-end training and inference on Raspberry Pi using real-world smart home data from five dementia households.

## II. Methods

### A. Data Collection

A total of nine participant–caregiver dyads were recruited for this work. Each dyad comprised a participant with mild-stage dementia and a co-resident caregiver, typically a partner. Unobtrusive ambient sensors recorded daily movement, along with temperature and light as additional channels for validating activities such as cooking and bathing as shown in Fig. 1. Activities were logged manually by the caregivers and standardized by an expert team in accordance with the Katz and IADL frameworks [6]. Of the nine homes, five produced usable datasets for the analysis reported here.

**Fig. 1.**
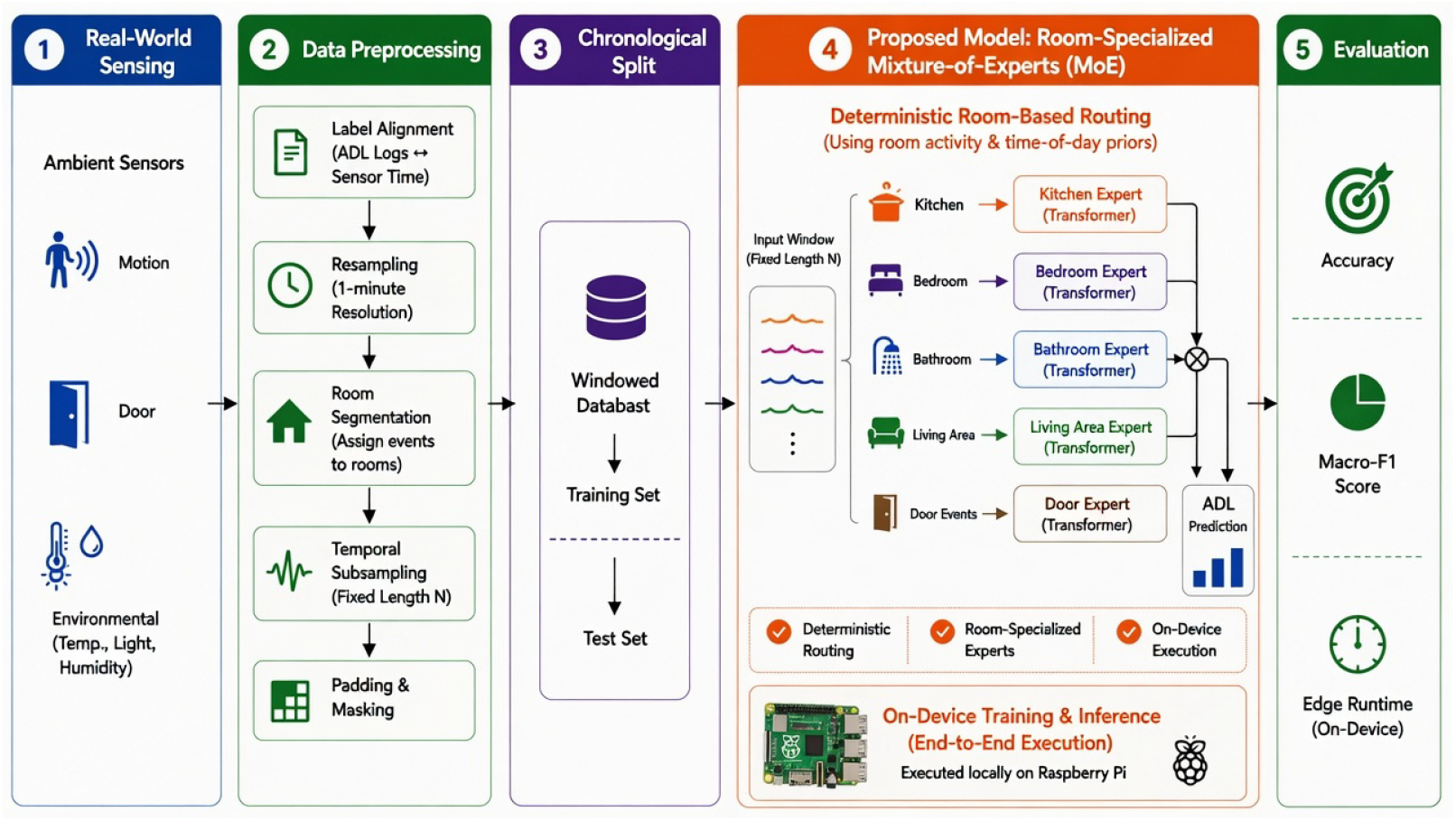
Overall workflow of the proposed method

**Fig. 2.**
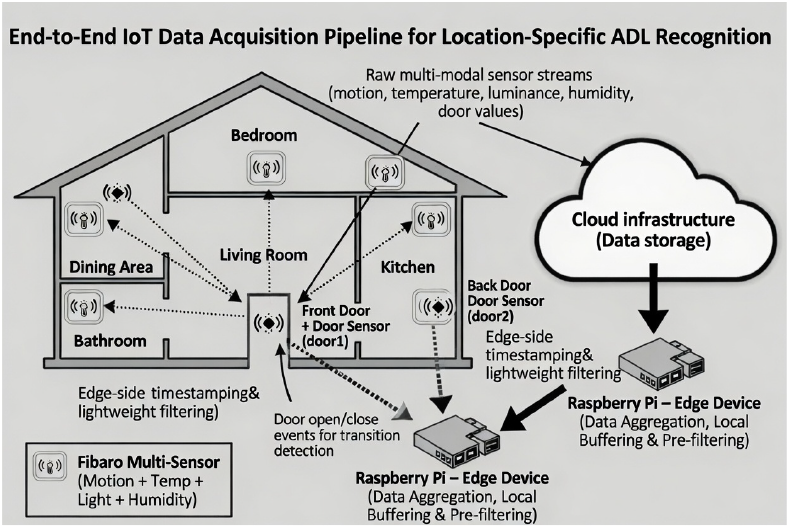
Smart home layout for end to end IoT data acquisition pipeline

### B. Data preprocessing

The collected ADL labels and sensor streams were resampled to a one-minute frequency [6]. Discrete channels such as motion and door sensors were forward-filled, while continuous channels, which are reported only on change, were reconstructed by spline interpolation between successive observations. The processed streams were combined by an inner join on timestamp to form a single unified dataset. This procedure was applied uniformly across all locations.

The resulting data is highly imbalanced across individual activities in both the training and test sets. Long-duration activities such as sleep dominate the data, which makes lower-frequency activities harder to recognize, especially when they occur in the same room as a dominant activity (for example, dressing and sleeping both occur in the bedroom). Rather than rebalancing the data with synthetic oversampling (e.g., SMOTE) or random undersampling, we adopt a subsampling strategy tied to room-based segmentation. Because each activity’s room is known from the participant-provided ground truth, the data is first segmented into room-based instances; for example, cooking, eating, and washing dishes are read from the kitchen sensors. Each contiguous activity interval, bounded by its ground-truth start and end times, forms one segment [6]. Each segment is then reduced to a fixed length of N ∈ 5, 30, 60, 240 time points before being passed to the model. When a segment is longer than N, we select N evenly-spaced points spanning its full duration; for example, with N = 60, a sleep segment lasting seven to eight hours is represented by 60 points sampled across the whole interval, preserving the temporal shape of the activity while bounding its size. Segments shorter than N are kept in full and zero-padded to length N, with the padded positions masked so that they contribute nothing to training or inference. The same procedure is applied to both the training and test sets.

This selective subsampling is what bounds the per-segment computational cost and makes on-device training feasible: regardless of an activity’s true duration, the model always processes at most N points. For each location, the last two days of data are held out as the test set and the remaining days are used for training.

### C. Design Rationale

Household activities naturally occur within specific rooms, resulting in localized sensor distributions (e.g., cooking predominantly observed in kitchens, while sleeping and dressing occur primarily in bedrooms). Learning these heterogeneous sensor distributions using a single global model increases both model complexity and computational requirements. Rather than learning all room-specific activity patterns jointly, we decompose the recognition task into independent room-specific experts. Each expert learns a smaller subset of activities from sensors associated with a single room, reducing the learning complexity while allowing each expert to specialize to a smaller activity space. Because the room associated with each activity segment is known from the sensor layout, routing can be performed deterministically without a learned gating network. This eliminates routing overhead while ensuring that only a single lightweight expert is executed during inference.

### D. Model Building

We implement the proposed architecture as a room-specialized Mixture-of-Experts (MoE) model composed of lightweight transformer experts [4]. Each expert consists of a lightweight transformer encoder specialized to the activity patterns of a single room. For each expert E, we employ a single-layer, two-head transformer encoder with a hidden dimension of 16. A deterministic, rule-based gate assigns every segment to exactly one expert using the room in which the activity occurs, so no learned routing network is required. Because the gate is deterministic, each expert is trained independently on the subset of data routed to it [5]. A fixed-length segment representation (length N), each expert processes a bounded input regardless of an activity’s true duration, enabling both end-to-end training and inference within the computational constraints of the Raspberry Pi.

### E. Inference testing

The whole training and testing has been done on the raspberry pi on the cpu option. All necessary libraries have been installed and the end to end training and testing was performed on cpu of pi and RTX 4090.

## III. Results

We evaluated the proposed room-specialized MoE on five dementia households to assess (1) recognition performance across homes and (2) the feasibility of end-to-end training and inference on Raspberry Pi.

Each household was modeled using a room-specialized MoE consisting of five room experts and one additional door expert for movement-related activities. Experts were automatically pruned when a corresponding room or activity was absent. Across the five households, the proposed framework achieved Macro-F1 scores ranging from 0.437 to 0.911 despite substantial differences in activity distributions and numbers of test segments (Table I). Performance varied across households, reflecting differences in activity distributions and the limited number of evaluation segments available at each site (7–27 segments). Macro-F1 was reported alongside accuracy because it better reflects performance under substantial class imbalance.

**TABLE 1.** Outcomes from Individual Sites at Window Length *L* = 5 Minutes.

| Location | $n_{\text{classifiers}}$ | $n_{\text{segments}}$ | Accuracy | Macro F1 |
| --- | --- | --- | --- | --- |
| Site 01 | 5 | 13 | 0.600 | 0.437 |
| Site 02 | 6 | 23 | 0.857 | 0.833 |
| Site 03 | 6 | 27 | 0.681 | 0.633 |
| Site 04 | 5 | 14 | 0.923 | 0.911 |
| Site 05 | 6 | 7 | 0.833 | 0.733 |

Table II compares per-class F1 across four segment lengths (N = 5, 30, 60, and 240). We evaluated multiple segment lengths to assess whether temporal subsampling preserved recognition performance while reducing computational cost. Recognition performance was generally stable across window lengths, although variability was observed for activities represented by very few evaluation segments, suggesting that temporal subsampling preserved sufficient activity information despite substantial reductions in input size. Several entries are based on only one or two evaluation segments (e.g., the uniform 1.00 scores for *Leisure Social* and the 0.00 scores for Transportation), and should therefore be interpreted as indicative rather than definitive performance estimates.

**TABLE 2.** Per-class F1 Score by Site Across Window Lengths (*N* = 5*/*30*/*60*/*240 minutes, respectively. “–” denotes the activity was absent at that site.)

| Activity | Site 01 | Site 02 | Site 03 | Site 04 | Site 05 |
| --- | --- | --- | --- | --- | --- |
| Eating | 0.50 / 0.50 / 0.67 / 0.67 | – / – / – / – | 0.50 / 0.50 / 0.29 / 0.67 | 1.00 / 1.00 / 1.00 / 1.00 | – / – / – / – |
| Dressing | 0.67 / 0.00 / 0.67 / 0.67 | 0.67 / 0.67 / 0.75 / 0.67 | – / – / – / – | 0.67 / 1.00 / 0.67 / 1.00 | – / – / – / – |
| Sleep_Rest | 0.67 / 0.67 / 0.67 / 0.67 | 0.50 / 0.50 / 0.80 / 0.50 | 1.00 / 1.00 / 0.00 / 0.00 | 0.80 / 1.00 / 0.80 / 1.00 | 1.00 / 1.00 / 1.00 / 1.00 |
| Leisure_Social | 1.00 / 1.00 / 1.00 / 1.00 | 1.00 / 1.00 / 1.00 / 1.00 | 1.00 / 1.00 / 1.00 / 1.00 | 1.00 / 1.00 / 1.00 / 1.00 | 1.00 / 1.00 / 1.00 / 1.00 |
| Transportation | 0.67 / 0.67 / 0.67 / 0.67 | 1.00 / 1.00 / 1.00 / 1.00 | 0.00 / 0.00 / 0.00 / 0.00 | – / – / – / – | 0.00 / 0.00 / 0.00 / 0.00 |

End-to-end training on the Raspberry Pi 4 CPU completed in approximately 16 minutes compared with 3 minutes on an RTX 4090 GPU. Although training on the edge device required approximately 5 times longer, the complete pipeline remained practical for on-device model updates without requiring cloud-based computation.

Together, these results demonstrate that room-specialized MoE enables practical transformer-based ADL recognition on resource-constrained edge devices while maintaining competitive recognition performance against the baseline performances [5].

## IV. Discussion

### A. Results from Subsampled data

We performed analysis using the subsampled datasets with the intention of running the end-to-end transformer based deep learning models on the edge computing devices such as raspberry pi 4. The data is selectively based on the window length. Across the evaluated window lengths (N = 5, 30, 60, and 240), recognition performance remained generally stable despite differences in activity distributions across households. Performance variability was largely associated with the limited number of evaluation segments available at some sites rather than the temporal compression itself. These findings suggest that the proposed subsampling strategy preserves sufficient temporal information for transformer-based ADL recognition while substantially reducing the computational burden for edge deployment. . Collectively, these results indicate that bounded temporal representations provide an effective trade-off between computational efficiency and recognition performance, making transformer-based ADL recognition practical on resource-constrained edge devices.

### B. Use of edge computing devices

The Raspberry Pi served not only as the smart home data acquisition platform but also as the deployment target for the proposed room-specialized MoE. This demonstrates that the same edge device used to continuously collect ambient sensor data can also support transformer-based ADL recognition without relying on cloud-based computation. Performing inference locally reduces the need to transmit continuous behavioral data outside the home, an important consideration for privacy-sensitive healthcare applications and regulatory compliance.

A key finding of this work is that transformer-based activity recognition can be redesigned to operate within the computational constraints of low-cost edge hardware. Rather than scaling model complexity to available computing resources, the proposed room-specialized MoE decomposes the recognition task into smaller room-specific experts while temporal subsampling bounds the computational cost of each activity segment. Together, these design choices enable end-to-end training and inference on a Raspberry Pi, suggesting that architectural simplification may be as important as hardware acceleration for practical edge AI deployment [8].

The ability to perform learning and inference directly on the edge device also has important implications for long-term smart home monitoring. As additional activity data become available, models can be updated locally without requiring continuous cloud connectivity or centralized storage of raw sensor streams. This edge-native workflow provides a foundation for privacy-preserving continual learning and federated learning frameworks, where model parameters rather than participant data are exchanged across homes [9].

Although training on the Raspberry Pi required more time than workstation-based training, the complete pipeline (i.e., preprocessing, model training, inference, and evaluation) was successfully executed on-device. This demonstrates that low-cost edge platforms can support practical AI-assisted monitoring for dementia care, particularly in settings where cloud infrastructure, high-performance computing resources, or reliable internet connectivity may be limited.

### C. Limitations

This study has several limitations. First, the proposed temporal subsampling strategy may discard fine-grained temporal transitions within long-duration activities. Although the overall recognition performance remained stable across different window lengths, more adaptive sampling strategies may better preserve informative temporal patterns while maintaining computational efficiency.

Second, the evaluation was conducted on five dementia households, with several activity classes represented by only a small number of evaluation segments. Larger and more diverse longitudinal datasets are needed to better assess the generalizability of the proposed architecture across homes, participants, and activity distributions.

Finally, this work focused primarily on demonstrating the feasibility of transformer-based ADL recognition on resource-constrained edge hardware. Additional systems-level evaluations, including memory utilization, inference latency, energy consumption, and long-term deployment reliability, remain important directions for future work.

## Data Availability

Data is currently not available online due to the agreements with the grant received from Alzheimer's Association

## Acknowledgment

All study procedures were reviewed and approved by the University of Missouri Institutional Review Board (IRB 2101666). This work is supported by Alzheimer’s Association Research Grant (24AARGD-NTF-1242722). The authors also like to acknowledge Sevara Pulatova, Abby Roessler, Claire Stam for their contribution to data collection.

## References

[1] Alzheimer’s Association, “2026 Alzheimer’s Disease Facts and Figures,” Chicago, IL, USA, 2026. [Online]. Available: https://www.alz.org/alzheimers-dementia/facts-figures

[2] National Alliance for Caregiving and AARP, Dementia Caregiving in the U.S., Bethesda, MD, USA: National Alliance for Caregiving, 2017. [Online]. Available: https://www.caregiving.org/wp-content/uploads/2020/05/Dementia-Caregiving-in-the-USFebruary−2017.pdf

[3] H. Liu and S. A. Fahmy, “Ask the Expert: Collaborative Inference for Vision Transformers with Near-Edge Accelerators,” arXiv preprint arXiv:2602.13334, Feb. 2026. [Online]. Available: https://arxiv.org/abs/2602.13334

[4] N. Shazeer, A. Mirhoseini, K. Maziarz, A. Davis, Q. Le, G. Hinton, and J. Dean, “Outrageously Large Neural Networks: The Sparsely-Gated Mixture-of-Experts Layer,” arXiv preprint arXiv:1701.06538, Jan. 2017, doi: 10.48550/arXiv.1701.06538. [Online]. Available: https://arxiv.org/abs/1701.06538

[5] V. R. Addepalli et al., “Room-Specialized Mixture-of-Experts for In-Home ADL Recognition with Ambient Sensors,” medRxiv, Jun. 2026, doi: 10.1101/2026.06.10.26355390. [Online]. Available: https://www.medrxiv.org/content/10.1101/2026.06.10.26355390v1

[6] S. Sutherland and S. Katz, “Concept mapping methodology: A catalyst for organizational learning,” Evaluation and Program Planning, vol. 28, no. 3, pp. 257–269, 2005, doi: 10.1016/j.evalprogplan.2005.04.017.

[7] A. E. Nieto-Vallejo, C. A. Parra-Rodriguez, and O. Ramirez-Perez, “Classification of Activities of Daily Living for Older Adults Using Machine Learning and Fixed Time Windowing Technique,” IEEE Sensors Journal, vol. 23, no. 24, pp. 31513–31522, Dec. 15, 2023, doi: 10.1109/JSEN.2023.3330630.

[8] Q. Song, S. Jing, S. Zhang, S. Zhang, and C. Huang, “Mixture-of-Experts for Distributed Edge Computing with Channel-Aware Gating Function,” arXiv preprint arXiv:2504.00819, Apr. 2025, doi: 10.48550/arXiv.2504.00819. [Online]. Available: https://arxiv.org/abs/2504.00819

[9] M. Ardakani, J. Malekar, and R. Zand, “LLMPi: Optimizing LLMs for High-Throughput on Raspberry Pi,” in Proc. IEEE/CVF Conf. Comput. Vis. Pattern Recognit. Workshops (CVPRW), Nashville, TN, USA, Jun. 2025, pp. 6433–6442, doi: 10.1109/CVPRW67362.2025.00634.

